# The Genetic Links to Anxiety and Depression (GLAD) Study: online recruitment into the largest recontactable study of depression and anxiety

**DOI:** 10.1101/19002022

**Authors:** Molly R. Davies, Gursharan Kalsi, Anamaria Brailean, Anthony J. Cleare, Jonathan R.I. Coleman, Charles J. Curtis, Susannah C.B. Curzons, Katrina A.S. Davis, Kimberley A. Goldsmith, Megan Hammond Bennett, Matthew Hotopf, Christopher Hüebel, Jennifer Leng, Bethany D. Mason, Monika McAtarsney-Kovacs, Dina Monssen, Elisavet Palaiologou, Carmine Pariante, Shivani Parikh, Alicia J. Peel, Katharine A. Rimes, Henry C. Rogers, Megan Skelton, Anna Spaul, Eddy L.A. Suarez, Bronte L. Sykes, Katie M. White, Allan H. Young, Evangelos Vassos, David Veale, Janet Wingrove, Thalia C. Eley, Gerome Breen

## Abstract

**Background:** Anxiety and depression are common, debilitating and costly. These disorders are influenced by multiple risk factors, from genes to psychological vulnerabilities and environmental stressors but research is hampered by a lack of sufficiently large comprehensive studies. We are recruiting 40,000 individuals with lifetime depression or anxiety, with broad assessment of risks to facilitate future research.

**Methods:** The Genetic Links to Anxiety and Depression (GLAD) Study (www.gladstudy.org.uk) recruits individuals with depression or anxiety into the NIHR Mental Health BioResource. Participants invited to join the study (via media campaigns) provide demographic, environmental and genetic data, and consent for medical record linkage and recontact.

**Results:** Online recruitment was effective; 41,892 consented and 26,877 participants completed the questionnaire by July 2019. Participants’ questionnaire data identified very high rates recurrent depression, severe anxiety and comorbidity. Participants reported high rates of treatment receipt. The age profile of sample is biased toward young adults, with higher recruitment of females and the better educated, especially at younger ages.

**Discussion:** This paper describes the study methodology and descriptive data for GLAD, which represents a large, recontactable resource that will enable future research into risks, outcomes and treatment for anxiety and depression.

**Highlights:**

- Online recruitment of 40,000 individuals with lifetime depression or anxiety (77 characters)
- Detailed online phenotyping combined with genetic and clinical data (66 characters)
- The study sample is severe, highly comorbid, with chronic psychopathology (62 characters)
- The study protocol enables recall of participants for future research and trials (82 characters)

The views expressed are those of the authors and not necessarily those of the NHS, NIHR, Department of Health or King’s College London

## Introduction

Anxiety and depression are the most common psychiatric disorders worldwide, with a lifetime prevalence of at least 30% (Bandelow & Michaelis, 2015; R. C. Kessler et al., 2005). They are the most common psychiatric disorders, are highly comorbid (Mineka, Watson, & Clark, 1998), and account for 10% of all years lived with disability (World Health Organisation, 2017). The World Health Organisation now considers depression to be the number one disorder by burden of disease (World Health Organisation, 2017). Many risk factors for depression and anxiety are shared, including psychological (e.g. cognitive biases (Michl, McLaughlin, Shepherd, & Nolen-Hoeksema, 2013; Naragon-Gainey, 2010)), environmental (e.g. stressful life events (Michl et al., 2013)), and genetic influences (which are ∼60-100% shared (Purves et al., 2017; Waszczuk, Zavos, Gregory, & Eley, 2014)). These findings on aetiology have been accompanied by an increasing evidence base of effective treatments, especially psychological (talking) therapies (Deacon & Abramowitz, 2004; Dobson, 1989; Hofmann & Smits, 2008). Nevertheless, despite advancements in psychological therapies as well as medication, clinicians are unable to predict which treatment will work for whom. This means that the choice of first and subsequent treatments currently progresses by trial and error at the cost of prolonged disability, reduced hope of success/engagement, and increased risk of adverse events.

Decades of work estimate twin study heritability of both anxiety and depression at ∼30-40% (Sullivan, Neale, & Kendler, 2000), rising to ∼60-70% for the recurrent forms of these disorders across several years (Kendler, Neale, R.C. Kessler, Heath, & Eaves, 1993). Recent UK Biobank analyses confirm that common genetic variants (single nucleotide polymorphisms, or “SNPs”) account for a smaller but still significant ∼15-30% of variation (“SNP-heritability”) in lifetime anxiety (Purves et al., 2017). Depression has a SNP-heritability of ∼12-14% and shows substantial genetic overlap with anxiety (Purves et al., 2017). Recent studies have identified 44 genetic variants for depression (Wray et al., 2018) and 5 genetic variants for anxiety (Purves et al., 2017), indicating that we are now entering the era where it will be possible to finally discover new biology for both disorders. In addition these genetic advances suggest that it may be time to include genetic risk factors in research aimed at developing new treatments, or predicting which therapies will work best for each patient (Keers et al., 2016; Lewis & Vassos, 2017).

Despite the broad range of established risk factors (Burcusa & Iacono, 2007; Craske et al., 2017; Fliege, Lee, Grimm, & Klapp, 2009), it is unclear how different types of influences combine to increase risk and influence treatment response. In particular, clinical/psychological (e.g. comorbidities), demographic (e.g. age), environmental (e.g. life events) and genetic influences have been studied largely independently, despite their evident interplay (Kendler, Gardner, & Prescott, 2002). As such, more multidisciplinary research is needed. However, efforts to better understand the aetiology of these disorders require large sample sizes and detailed information on symptom presentation. Many studies exist worldwide with either the number of participants or the thorough phenotyping needed, but few have both the scale and detail required. In response to these issues, the Genetic Links to Anxiety and Depression (GLAD) Study was developed to recruit 40,000 individuals into the newly established National Institute for Health Research (NIHR) Mental Health BioResource. This will be one of the largest single studies to date (as compared to previous meta-analyses) of individuals with a lifetime experience of clinical depression and anxiety.

The NIHR Mental Health BioResource is an integral part of the overall NIHR BioResource for Translational Research (https://bioresource.nihr.ac.uk/), which has previously recruited approximately 100,000 individuals. However, a large majority of volunteers are either healthy controls or have a physical health condition and no reported mental health disorder, which led to the creation of the NIHR Mental Health Bioresource. GLAD is its first project and has the specific goal of recruiting a large number of participants with anxiety and depression to facilitate future recall and secondary data analysis studies. Recall studies would involve recontacting participants to take part in further research, such as clinical trials aimed at developing better therapies and interventions for anxiety and depression. The GLAD Study will also directly explore both genetic and environmental risk factors for depression and anxiety disorders, including the potential of polygenic risk scores (Purcell et al., 2009) created from analyses of related phenotypes to predict response to treatment and prognosis. GLAD aims to facilitate studies which need to combine genetic, phenotypic, and environmental exposure data by creating a large, homogeneously phenotyped cohort of individuals with depression or anxiety with all these types of data.

## Methods

### Pre-Launch Preparation

A focused and intense social media campaign was utilised to inform the public about the GLAD study. We hoped that individuals with anxiety and depression would be encouraged to join given the online methodology, allowing them to respond to the questionnaires in a convenient time and location. The timeline leading up to the media launch is outlined in Figure 1. Experts in the field, patients and individuals with lived experience of depression or anxiety were consulted at each stage of study development. The study team also consulted regularly with collaborators from the Australian Genetics of Depression Study who provided guidance and advice based on their experience of successfully implementing a similar study design in Australia (https://www.geneticsofdepression.org.au/).

**Figure 1.**
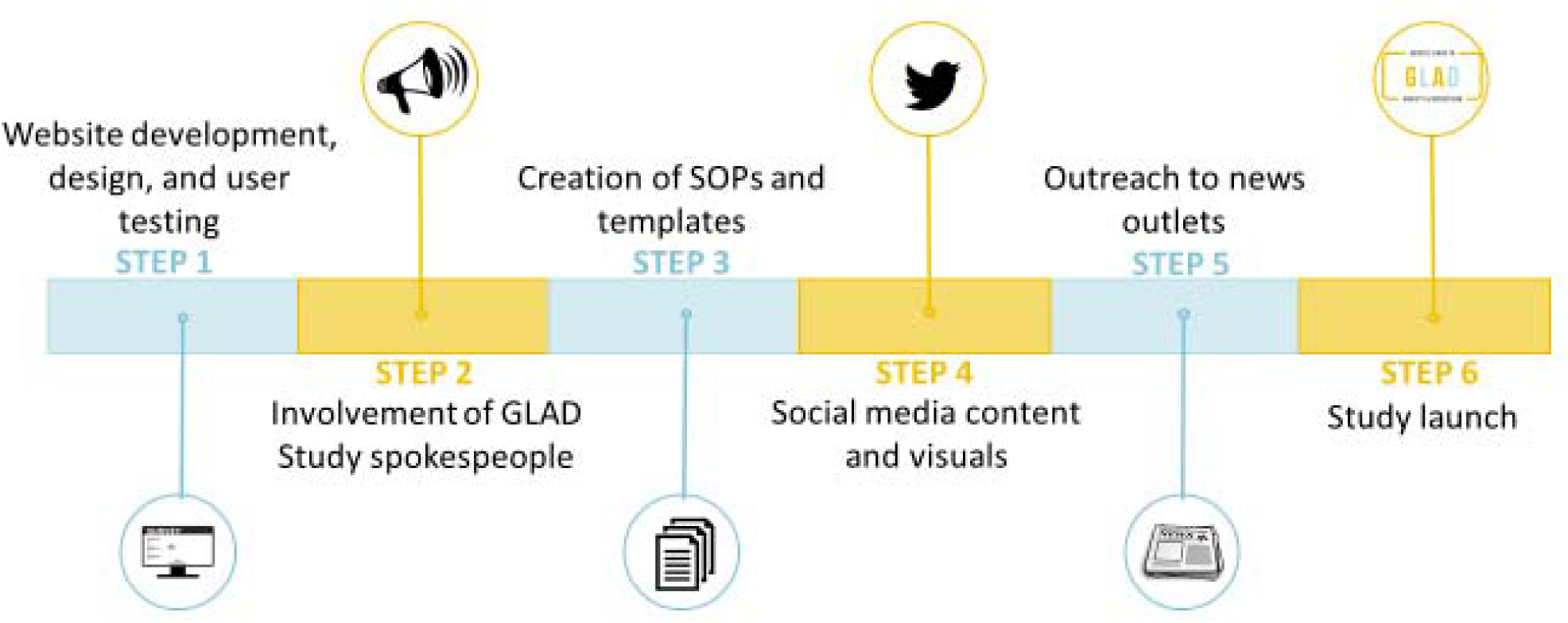
Steps to prepare for launch of the GLAD Study

#### Website Development, Design, and User Testing

The website was developed in collaboration with a local company (Mindwave Ventures) in various stages. During the early stages of development and the design phase, we conducted user testing on the website by contacting individuals with lived experience through various routes. The first user testers were patient volunteers within the Improving Access to Psychological Therapies (IAPT) service at South London and Maudsley (SLaM) National Health Service (NHS) Foundation Trust. The second group of volunteers were members of the King’s College London Service User Research Enterprise (SURE) and the Feasibility and Acceptability Support Team for Researchers (FAST-R). Third were volunteer individuals with lived experience who were actively involved with the charity Mind. Finally, staff from the charities Mind, MQ Mental Health, and the Mental Health Foundation gave feedback. All individuals provided input on the website content and usability, appearance, and study information and all volunteers (but not charity staff) were compensated £10 for their time. All feedback was reviewed by the study team and the lead investigators and incorporated into the final version of the website.

#### GLAD Study Spokespeople

A wide range of charities, professional bodies, companies, influencers (defined as individuals with a large number of followers on social media), and individuals with lived experience were identified based on previous advocacy, involvement, or openness of mental health and treatment. These individuals and organisations were approached by the public relations (PR) company (Four Health Communications), the study team, and/or the co-investigators to introduce the study and invite them to collaborate.

We received support from many UK charities including Mind, MQ, Mental Health Foundation, Anxiety UK, OCD Action, Bipolar UK, HERO, No Panic, the Charlie Waller Memorial Trust, BDD Foundation, Maternal OCD, Rethink Mental Illness, SANE, MHFA England, Universities UK, and UK Youth. We also received significant support from two UK professional bodies for psychologists and psychiatrists, the British Psychological Society and the Royal College of Psychiatrists. Finally, three organisations (Royal Mail, Priory Group, and Barnardo’s) agreed to circulate study information internally to their employees, and Priory Group additionally shared study information on social media.

Many of the charities, professional bodies, and influencers provided a quote about the GLAD Study to be shared with news outlets and on social media. Other forms of support from charities or professional bodies included circulating study information in member newsletters or magazines, publishing study information on their websites, and sharing the study on social media (http://wke.lt/w/s/Ymfhz). Influencers helped to promote the study on their social media channels or blogs. Individuals with lived experience allowed the GLAD Study team to share their stories on social media and to news outlets. Of utmost importance, these individuals were available for interviews with broadcast and newspaper agencies and gave a personal voice to the campaign. Spokespeople who became actively involved in the publicity and circulation of the GLAD Study in the media and/or online were vital to the success of the campaign.

#### Standard Operating Procedures and Templates

We created a variety of standard operating procedures (SOPs) and response templates to prepare for the logistical and administrative aspects of study launch and management. These included SOPs for saliva sample kit preparation and posting, website management, data protection, and participant contact. We established guides and SOPs for responding to participant questions, concerns, and expressions of distress. These response guidelines were reviewed by clinicians and by the Mental Health Foundation to ensure responses were appropriate, straightforward, and clear.

#### Social Media Content and Visuals

Social media content and visuals were designed with the aid of the PR company (which had experience of working on NHS projects), an independent videographer, and an animation company. These included a range of infographics and short videos, each designed to be informative and provide basic details for the public about aims of GLAD and how to join the project. The campaign was targeted to a younger demographic (age 16 - 30) to enable more prolonged collection of longitudinal data and reflect the young age of onset of depression and anxiety disorders (Bandelow & Michaelis, 2015; Christie et al., 1988; R. C. Kessler et al., 2005).

The infographics (Figure 2) emphasised the simplicity and ease of taking part. The consent animation (https://youtu.be.com/SAE0yVNvWaA) outlined key elements of the consent process to provide a simple and clear way to learn about the study, consistent with guidance from the Health Research Authority regarding multimedia use as a patient-friendly method of delivering vital consent information (Health Research Authority, 2017). The consent animation was also included with the website version of the information sheet. The second video was a short film (https://youtu.be/wzgvS8gU2Ss which outlined the study sign-up steps in a real-life, relatable way. Finally, the animation (https://youtu.be.com/HUI5eFXevvk) was an engaging ‘call to action’ to join the community of volunteers participating in the GLAD Study.

**Figure 2.**
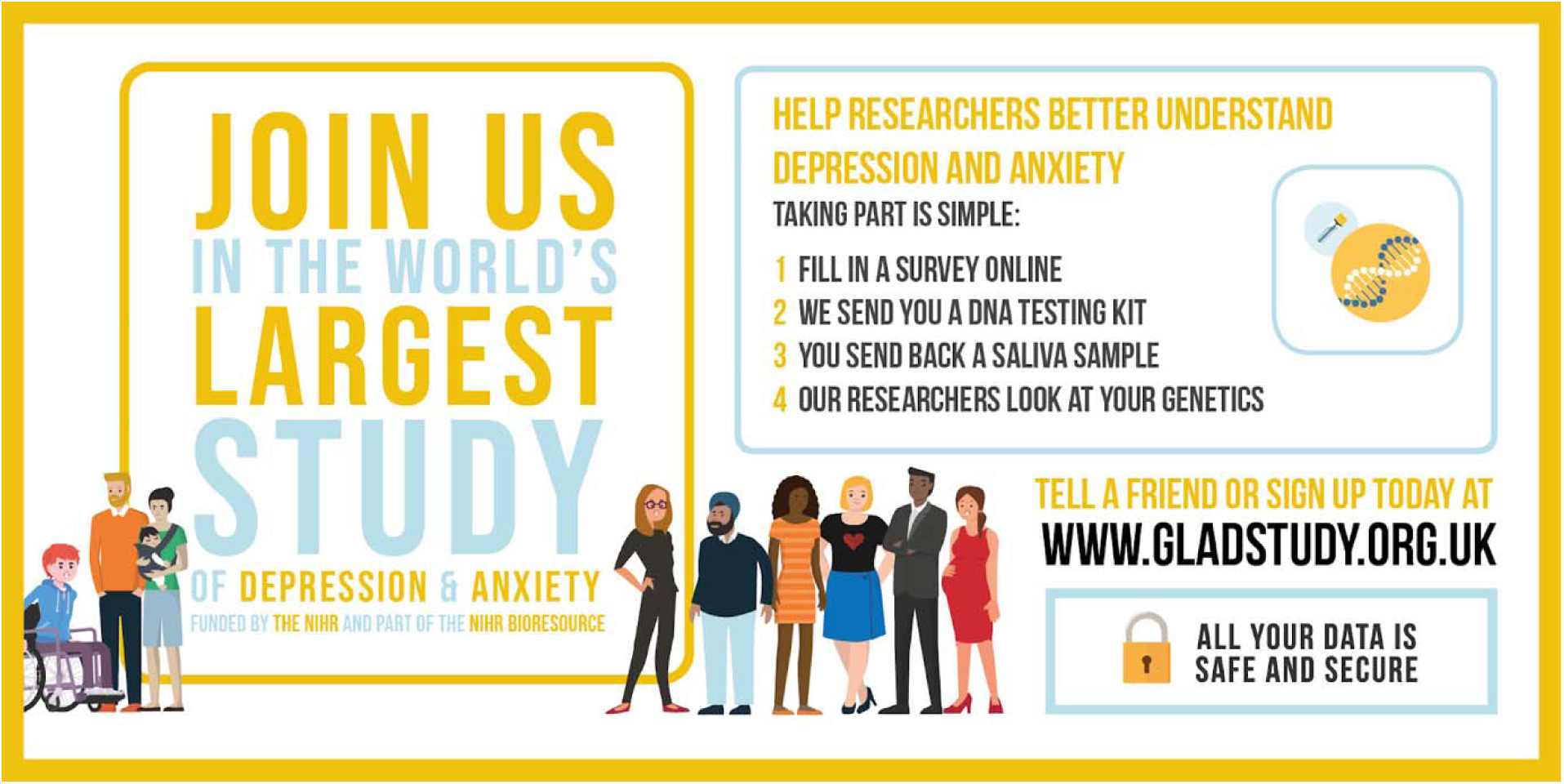
GLAD Study infographic shared on Twitter Example of the infographic shared on Twitter to promote the GLAD study and explain the key steps involved in signing up

We created a 6-week social media schedule for planned posts across Facebook, Instagram, and Twitter platforms which included both organic and promoted content. Most of the posts were accompanied by a video, infographic, or other imagery developed by the study team.

#### Outreach to News Outlets

The PR company circulated the press release and study information to news and broadcast agencies across England. Interested parties organised interviews with the study investigators and/or influencers, charity or professional body representatives, and individuals with lived experience. The press releases and broadcasts were embargoed until the launch date to prevent pre-emptive publicity and provide uniform, widespread coverage across different media.

### Recruitment

#### Media Campaign: Study launch

The PR company helped us to organise a widespread national media campaign which included traditional media (TV, radio, newspaper) and social media (Twitter, Facebook, Instagram) outreach during the study launch. Examples from the campaign can be viewed at this link: http://wke.lt/w/s/Ymfhz. In the first 24 hours, 8004 participants had registered to the website, demonstrating the effectiveness of this strategy. The prepared social media schedule helped to maintain our recruitment numbers over the 6-week period. Between 300-1,500 participants signed up to the study daily, with numbers fluctuating based on campaign expenditure and social media support from influencers and organisations.

#### Clinical Recruitment

In the 9 months following the launch, many NHS organisations contacted us interested in supporting the GLAD Study. As of July 2019, these included 12 Clinical Research Networks (CRN), 7 family doctor/general practitioner (GP) practices, 93 Trusts, 1 Clinical Commissioning Group (CCG), and 1 GP Federation. Sites were classified in 2 different categories with varying levels of involvement: 1) advertising sites which displayed study posters and leaflets in clinics and waiting areas, and 2) recruiting sites which conducted mail-outs, approached patients in the clinic or over the phone, and assisted patients through the study sign-up process.

### Sign-up Process

The sign-up process for enrolment in GLAD is given in Figure 3. Personal information and phenotypic data are collected entirely online through the GLAD Study website (www.gladstudy.org.uk). Participants register on the website with their name, email address, phone number, date of birth, sex, and gender. They are then able to read the information sheet and provide consent. As part of the consenting process participants agree to long-term storage of their sample, requests to complete follow-up questionnaires, anonymised data sharing, recontact for future research studies based on their phenotype/genotype information, and access to their full medical and health related records. Following consent, participants complete the sign-up questionnaire to assess their eligibility. Eligibility criteria are restricted to participants that meet DSM-5 criteria for MDD or any anxiety disorder, based on responses to screening measures in our online sign-up questionnaire. More information about the measures in the sign-up questionnaire are included below. Eligible participants are then sent an Isohelix saliva DNA sample kit. Saliva samples are sent via Freepost to the NIHR National Biosample Centre in Milton Keynes for processing and storage. Once participants have returned their saliva sample, they become full members of the GLAD Study (and of the NIHR BioResource).

**Figure 3.**
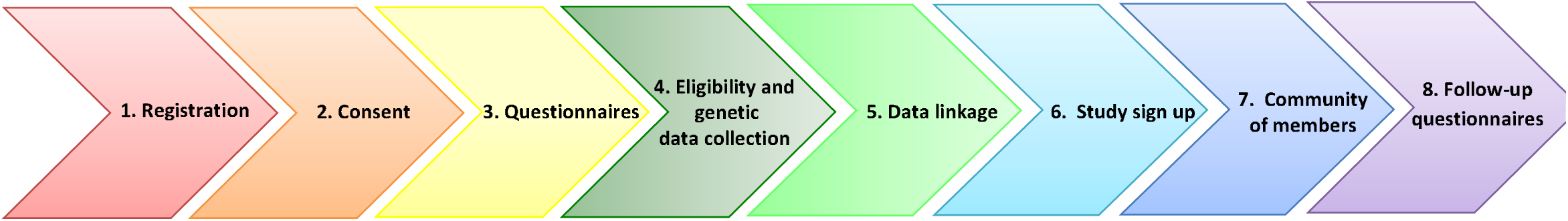
Stages of participant sign-up and involvement in the GLAD Study

Email reminders are sent to participants who do not complete their sign-up steps. Up to four automated reminder emails are sent through the website up to 6 months post starting sign-up. Once online sign-up steps are completed, a maximum of four reminder emails are sent to participants regarding the saliva samples up to 6 months from the date the kit was sent. We conduct additional phone call reminders to participants who have not returned their saliva kits within three months of signing-up.

Clinical data will be linked to genotype and phenotype data to provide additional information about participants’ medical history, diagnoses, and treatments relevant to current and future research projects. Eligibility for collaborating studies can then be assessed utilising all phenotypic, genetic, and clinical data. Members may then be contacted up to four times per year to take part in future studies, either through the website or by the GLAD Study or NIHR BioResource teams, with access granted via established NIHR BioResource access protocols (see below for further detail). As part of the community of members, participants will also have access to useful information and links on the “Useful Links” page of the study website. Members will also be invited to take part in follow-up questionnaires to provide longitudinal data on their symptoms.

### Measures

#### Sign-up Questionnaire

The sign-up questionnaire was designed to assess core demographic and personal information as well as detailed psychological and behavioural phenotyping relevant to anxiety and depression (for full measure names see Table 1). We included lifetime measures of major depressive disorder, atypical depression, and generalised anxiety disorder (adapted from CIDI-SF (Patten, Brandon-Christie, Devji, & Sedmak, 2000) and ICD-11 checklists (Kogan et al., 2016)), supplemented with items enabling lifetime assessment of DSM-5 specific phobia, social phobia, panic disorder and agoraphobia. These items were adapted from the Australian Genetics of Depression Study questionnaire (https://www.geneticsofdepression.org.au/).

**Table 1.** GLAD Study sign-up questionnaire measures

| Assessment/Topic | Purpose | Source |
| --- | --- | --- |
| **Lifetime depression** | Lifetime assessment of major depressive disorder | Adapted Composite International Diagnostic Interview – Short Form (CIDI-SF) |
| **Lifetime atypical depression** | Lifetime assessment of atypical depression | Adapted DSM-5/ICD-11 checklists |
| **Lifetime anxiety** | Lifetime assessment of generalised anxiety disorder | Adapted Composite International Diagnostic Interview – Short Form (CIDI-SF) |
| **Lifetime specific phobia** | Lifetime assessment of specific phobia | Adapted DSM-5/ICD-11 checklists |
| **Lifetime social phobia** | Lifetime assessment of social phobia | Adapted DSM-5/ICD-11 checklists |
| **Lifetime panic disorder** | Lifetime assessment of panic disorder | Adapted DSM-5/ICD-11 checklists |
| **Lifetime agoraphobia** | Lifetime assessment of agoraphobia | Adapted DSM-5/ICD-11 checklists |
| **Current depressive symptoms** | Assess severity and presence/absence of current depression. | Patient Health Questionnaire 9-items (PHQ-9) |
| **Treatment-resistant depression** | Assess treatments during most recent or current depressive episode to screen for treatment-resistant | Maudsley Staging Model of Treatment Resistance, Modified Self-Report (MSM-SR) |
| **Possible mania/hypomania** | Screen participants for possible episodes of hypomania or mania | The Mood Disorder Questionnaire (MDQ) |
| **Current general anxiety symptoms** | Assess the severity and presence/absence of current anxiety | Generalised Anxiety Disorder 7-item scale (GAD-7) |
| **Current post-traumatic stress disorder symptoms** | Screen for presence and severity of PTSD symptoms. | Post-traumatic stress disorder checklist, civilian version 6-items (PCL-6) |
| **Alcohol use** | Screen for alcohol dependence in the past year | Alcohol Use Disorders Identification Test (AUDIT) |
| **Psychotic symptoms** | Assess lifetime psychotic experiences | Adapted CIDI (McGrath et al., 2015) |
| **Personality disorders** | Assess the severity of personality disorders. | Standardized Assessment of Severity of Personality Disorder 9-items (SASPD) |
| **Work and social adjustment** | Assess functional impairment due to psychopathology | Work and Social Adjustment Scale (WSAS) |
| **Subjective wellbeing** | Assesses subjective well-being, specifically positive emotion and meaning | Two hedonic ('positive emotion') questions and a eudaimonic ('meaning') question from WHO-Quality of Life (WHOQOL) |
| **Adversities in childhood** | Identify presence/absence of childhood abuse and neglect | Childhood Trauma Screener short version (CTS-5) |
| **Adult domestic abuse and catastrophic trauma** | Identify victims of adult domestic abuse. | A 5-item domestic abuse screen developed for the UK Biobank Mental Health Questionnaire (Davis et al., 2018), adapted from the national crime survey and CTS-5, and 5 items on catastrophic trauma |

We also assessed current depressive symptoms (PHQ-9 (Kroenke, Spitzer, & Williams, 2001)) and those with a score of 5 or above on the PHQ 9 (indicating a current episode of depression) were asked additional questions related to current treatment-resistant depression (MSM-SR (Fekadu, Donocik, & Cleare, 2018)). Other measures of current psychopathology included possible mania/hypomania (MDQ (Hirschfeld, 2002)), current general anxiety symptoms (GAD-7 (Spitzer, Kroenke, Williams, & Löwe, 2006)), post-traumatic stress disorder symptoms (PCL-6 (Lang et al., 2012)), alcohol use (AUDIT-C (Saunders, Aasland, Babor, de La Fuente, & Grant, 1993)), psychotic symptoms (adapted CIDI (McGrath et al., 2015)), personality disorder symptoms (SAS-PD (Olajide et al., 2018)), and work and social adjustment (WSAS (Mundt, Marks, Shear, & Greist, 2002)). Additional measures were included to assess subjective well-being (Group, Whoqol, 1994), recent adverse life events, 5 childhood trauma items (Glaesmer et al., 2013) representing the 5 subscales of the full Childhood Trauma Questionnaire (Pennebaker & Susman, 2013), domestic violence (Davis et al., 2018), and catastrophic trauma (Davis et al., 2018).

To facilitate future meta-analyses with other cohorts, measures were selected when possible to coincide with the UK Biobank Mental Health Questionnaire (MHQ) (Davis et al., 2018). Some aspects of the UK Biobank MHQ were not selected for the sign-up questionnaire in order to make space for additional measures more relevant here. Furthermore, detailed questions on self-harm and suicide were asked during sign-up due to concerns from the study team and the SLaM Research and Development (R&D) department of insufficient clinical support in the case of reported adverse events, although the single suicidal ideation item in the PHQ-9 was retained.

#### Optional Questionnaires

Once participants completed the sign-up questionnaire, they were invited to take part in additional, optional questionnaires (SM 1). Optional questionnaires to assess a wider number of psychiatric phenotypes and symptoms included measures of fear (Fear Survey Schedule (Wolpe & Lang, 1964)), drug use (DUDIT (Berman, Bergman, Palmstierna, & Schlyter, 2003)), obsessive-compulsive disorder (OCI-R (Foa et al., 2002)), post-traumatic stress disorder (PCL-5 (Frank W. Weathers et al., 2013)), trauma (K Kopenen, personal communication), postnatal depression (EPDS (Cox, Holden, & Sagovsky, 1987)), body dysmorphic disorder (DCQ (Mancuso, Knoesen, & Castle, 2010)), eating disorders (Allison, Stunkard, & Thier, 2004; Herman et al., 2016; Hildebrandt, Langenbucher, & Schlundt, 2004; Thornton et al., 2018), and vomit phobia (SPOVI (Veale et al., 2013)).

Other optional projects relate to lifestyle and personal history, asking detailed questions on participants’ experience of healthcare, life events (F. W. Weathers et al., 2013), work and sleep, general health and lifestyle, gambling, and headaches and migraines (https://www.geneticsofdepression.org.au/). Optional questionnaires were made available to all participants, except those on vomit phobia and postnatal depression which were only offered to participants based on responses to screening questions in the sign-up questionnaire. Additional follow-up questionnaires will be sent to participants annually to provide longitudinal data on symptoms and outcomes.

### Genotyping

All GLAD samples will be genotyped as part of core NIHR BioResource funding. For genotyping we are using the UK Biobank v2 Axiom array, consisting of >850,000 genetic variants (Bycroft et al., 2018), designed to give optimal information about other correlated genetic variants. This careful design means that imputation of the data with large whole genome sequencing reference datasets will yield >10 million common genetic variants per individual. We use Affymetrix software and the UK Biobank pipeline software to assign genotypes and perform standard quality control measures in PLINK (Chang et al., 2015) or equivalent software packages and R (Team, 2015). Analyses will be conducted in PLINK in the first instance.

### Ethical Approval

The GLAD Study was approved by the London - Fulham Research Ethics Committee on 21st August, 2018 (REC reference: 18/LO/1218) following a full review by the committee. The NIHR BioResource has been approved as a Research Tissue Bank by the East of England - Cambridge Central Committee (REC reference: 17/EE/0025). Prior to submission for ethical approval, this research was reviewed by a team with experience of mental health problems and their carers who have been specially trained to advise on research proposals and documentation through the Feasibility and Acceptability Support Team for Researchers (FAST-R): a free, confidential service in England provided by the NIHR Maudsley Biomedical Research Centre via King’s College London and South London and Maudsley NHS Foundation Trust.

### Analysis

#### Genetics and combined predictors of anxiety and depression

The GLAD data can be used alongside UK Biobank for meta-analyses, with UK Biobank participants and/or other NIHR Bioresource participants as healthy controls, to provide adequate statistical power. One of our primary genomic aims is to utilise polygenic scores created from very large genome-wide analyses of related traits (e.g. anxiety) as potential predictors of depression, anxiety, and treatment response in our sample. There are a number of well-powered polygenic scores now available in the field not only for psychiatric traits but also intelligence and its proxies(Selzam et al., n.d.) and other relevant predictors. We aim to combine these polygenic scores with significant clinical predictors to produce a combined clinical/genetic risk index.

We envision the GLAD sample to also be incorporated either in large genome-wide association studies, contributing to meta-analyses, or in clinical trials, for example researching genetic, epidemiological or social risk factors of anxiety and depression.

### Future Research and Collaborations

Researchers wishing to access GLAD Study participants or data are invited to submit a data and sample access request to the NIHR BioResource to request a collaboration, following the procedures outlined in the access request protocol (SM 2). Applications will be reviewed by the NIHR Steering Committee to assess the study aims and to check ethical approvals and protocols. Collaborations can range from sharing of anonymised data or samples to recontact of participants for additional studies, including experimental medicine studies and clinical trials. Eligibility for future research can be targeted to specific genotypes and/or phenotypes of interest.

## Results

### Sample Descriptors

Recruitment of the GLAD Study is ongoing. All results that follow are from participants recruited before 04 July 2019. As of this date, 41,892 participants had consented to the GLAD Study, with approximately 60% drop-off rate at each stage of the sign-up process. This resulted in 27,475 (65.6%) having completed the questionnaire, with 27,264 (99.2%) screened as eligible and sent a saliva kit. Of those participants which were sent a kit, 17,539 (64.3%) have returned a saliva sample thus far.

In the current sample, the mean age is 36.9 with most participants being female, white, and in paid employment or self-employed (Table 2). The GLAD sample is younger and more female (Figure 4) and has a higher proportion of individuals with a university degree (Figure 5) than the general population of England. For example, of those aged 25-29 years, 15.2% of our sample is female compared to only 4.5% of the population of England. This difference is seen across most/all age-groups. Similarly, in the age-range 16-24 years, 40.4% of the sample have a university degree whereas across England as a whole this is only 13.7%.

**Table 2.** Descriptors of GLAD sample, July 2019

|  | Percent |
| --- | --- |
| Sex |  |
| Male | 22.8% |
| Female | 76.5% |
| Gender |  |
| Transgender | 1.4% |
| Sexuality |  |
| Heterosexual | 72.4% |
| Homosexual | 6.3% |
| Bisexual | 15.8% |
| Asexual | 1.3% |
| Self-define | 2.8% |
| Ethnicity |  |
| White | 93.0% |
| Mixed | 2.9% |
| Asian or Asian British | 1.4% |
| Black or Black British | 0.4% |
| Arab | 0.1% |
| Other | 1.1% |
| Education (highest level achieved) |  |
| College or university degree | 59.6% |
| A levels/AS levels or equivalent | 21.5% |
| O levels/GCSEs or equivalent | 12.5% |
| CSEs or equivalent | 1.4% |
| Employment |  |
| Employed | 61.7% |
| Retired | 5.4% |
| Unable to work because of sickness or disability | 9.4% |
| Unemployed, looking after home/family, or doing voluntary work | 8.7% |
| Full or part-time student | 13.1% |
| Relationship status |  |
| Single | 27.4% |
| Relationship | 32.0% |
| Married/civil partnership | 31.5% |
| Divorced/separated | 6.7% |
| Widowed | 0.8% |
| Smoking status |  |
| Current | 16.0% |
| Former | 31.2% |
| Never | 52.1% |
| Handedness |  |
| Right handed | 87.4% |
| Left handed | 11.0% |
| Ambidextrous | 1.6% |
| Received treatment* |  |
| Any treatment | 95.2% |
| Psychological (talking) therapies | 81.7% |
| Medication | 88.5% |

*\*Treatment percentages were only calculated for participants who screened positively for MDD or GAD. Participants were asked to select all that applied in response to the question "Did you ever try or are currently trying the following for these problems?" Participants are considered to have received treatment if they indicated they tried medication prescribed for at least two weeks or tried psychotherapy or other talking therapy more than once.*

**Figure 4.**
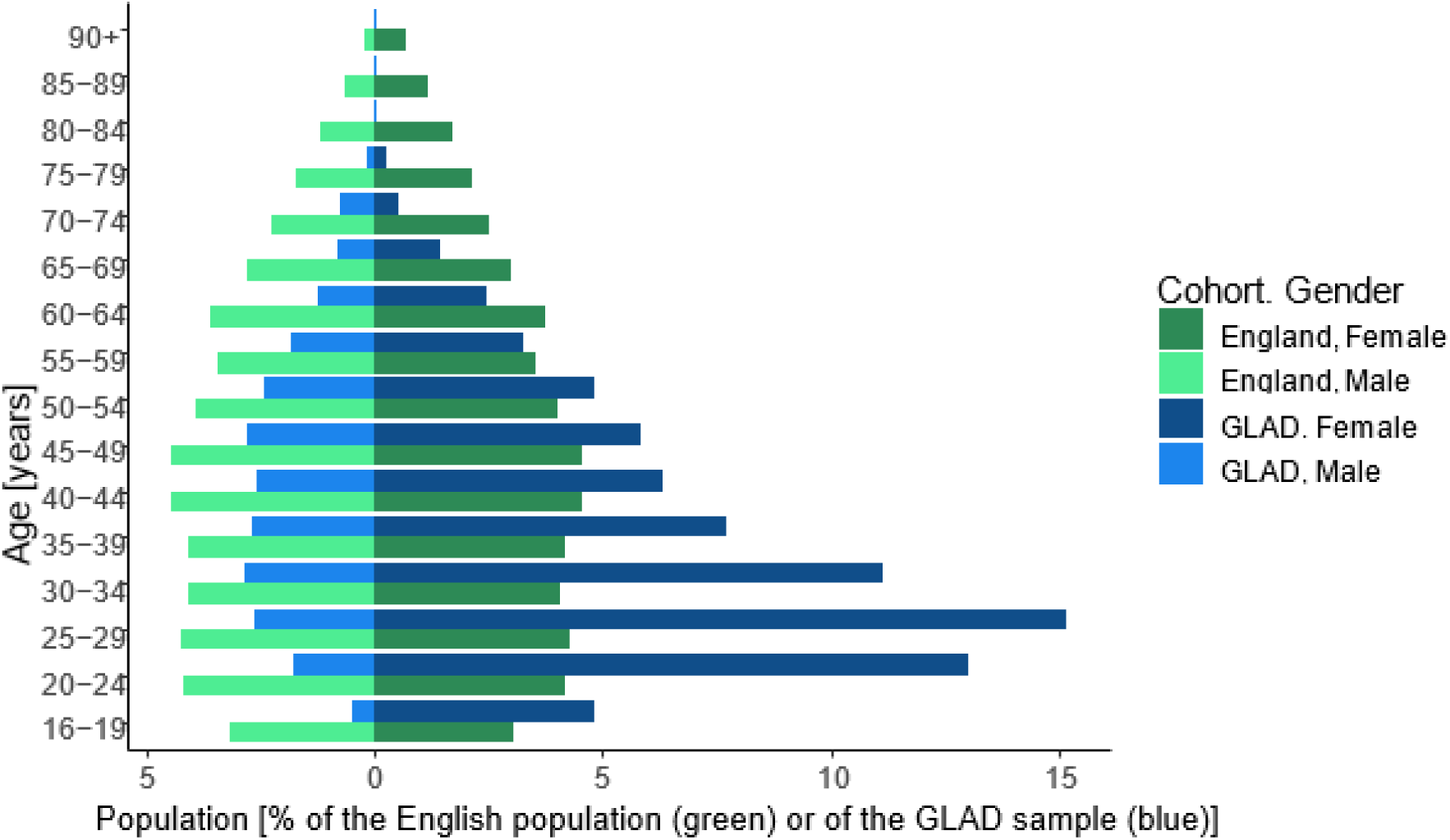
Distribution of gender and age in the GLAD sample compared to the England population Comparison of age and sex characteristics between the GLAD sample and the general population of England as measured in the 2011 census (Office for National Statistics, National Records of Scotland, & Northern Ireland Statistics and Research Agency, 2016). Proportions of males in each age group are displayed on the left side of the graph, with female proportions on the right. GLAD participants are represented in light (male) and dark (female) shades of blue, while the England population is represented in light (male) and dark (female) shades of green.

**Figure 5.**
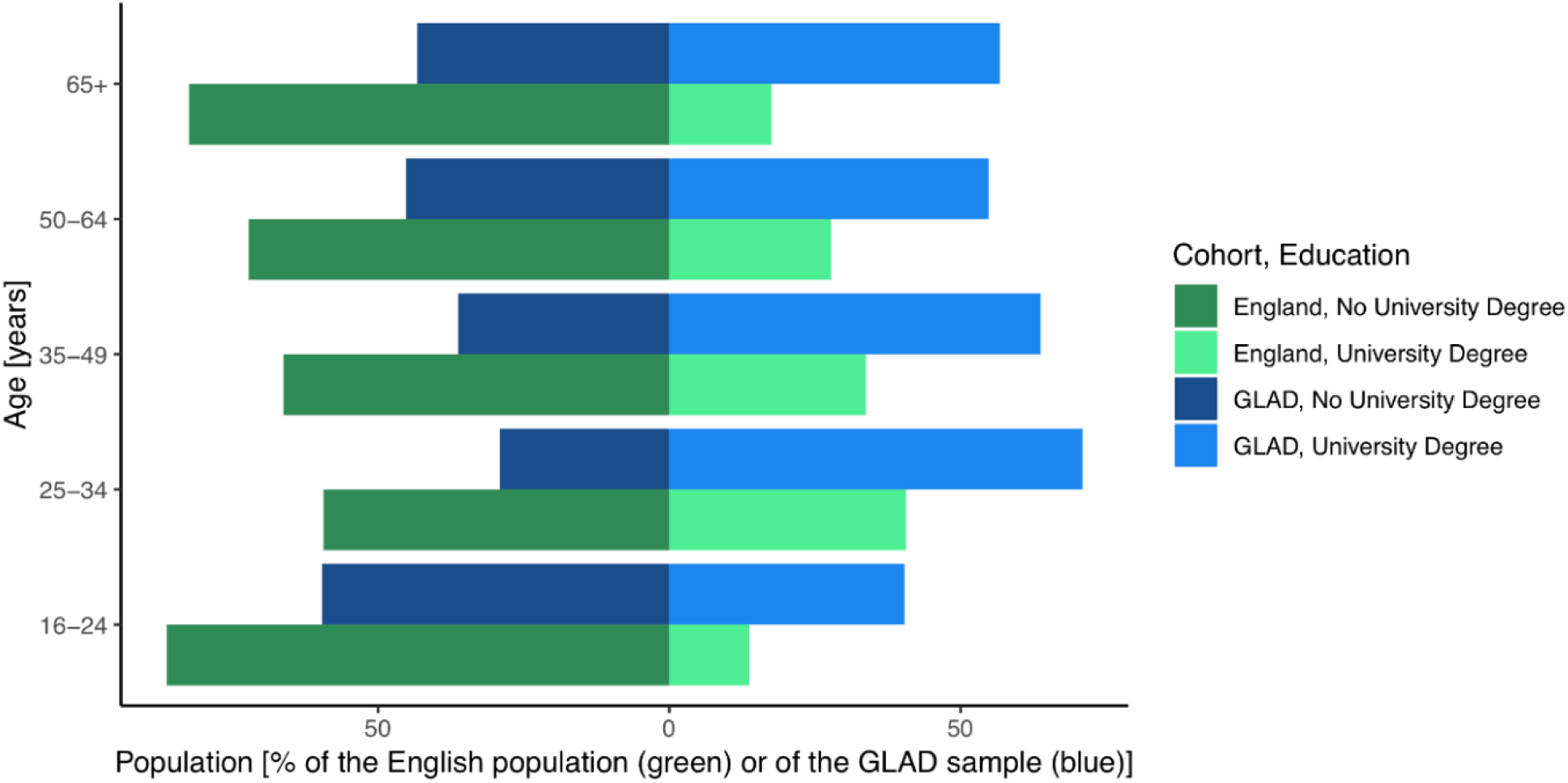
Proportion of individuals with university degrees by age group in the GLAD sample compared to the England population Comparison of education level by age group between the GLAD sample and the general population of England as measured in the 2011 census (Office for National Statistics et al., 2016). Proportions of individuals without university degrees in each age group are displayed on the left side of the graph, with proportions of individuals with university degrees on the right. GLAD participants are represented in dark (no university degree) and light (university degree) shades of blue, while the England population is represented in dark (female) shades of green and light (university degree).

A high proportion of GLAD participants report the occurrence of at least one form of trauma or abuse throughout their lives. Child abuse was most commonly reported, with emotional abuse by parents or family being endorsed by almost 40% of the sample. Other forms of life stress are also frequently endorsed with 82.8% of the sample reporting at least one traumatic experience in their lives. Of note, few participants reported periods of inability to pay rent, reflecting low rates of poverty in the sample.

### Psychopathology

As shown in Figure 6, the majority of participants reached diagnostic criteria for major depressive disorder (MDD), followed by panic disorder and generalised anxiety disorder (GAD). The majority of participants with depression reported recurrent episodes across the lifespan. A figure demonstrating self-reported clinician-provided diagnoses of mental health disorders (as opposed to questionnaire-assessed) in the GLAD sample can be found in supplementary materials (SM 3). Of note, self-reported clinician-provided diagnoses of GAD (77.3%) are twice as high as cases assessed by the questionnaire. The sample also has high rates of comorbidity, with 66.2% of screened MDD cases also screening positively for at least one of the anxiety disorders (generalised anxiety, specific phobia, social phobia, panic disorder, or agoraphobia), and 92.4% of screened GAD cases also screening positively for major depressive disorder.

**Figure 6.**
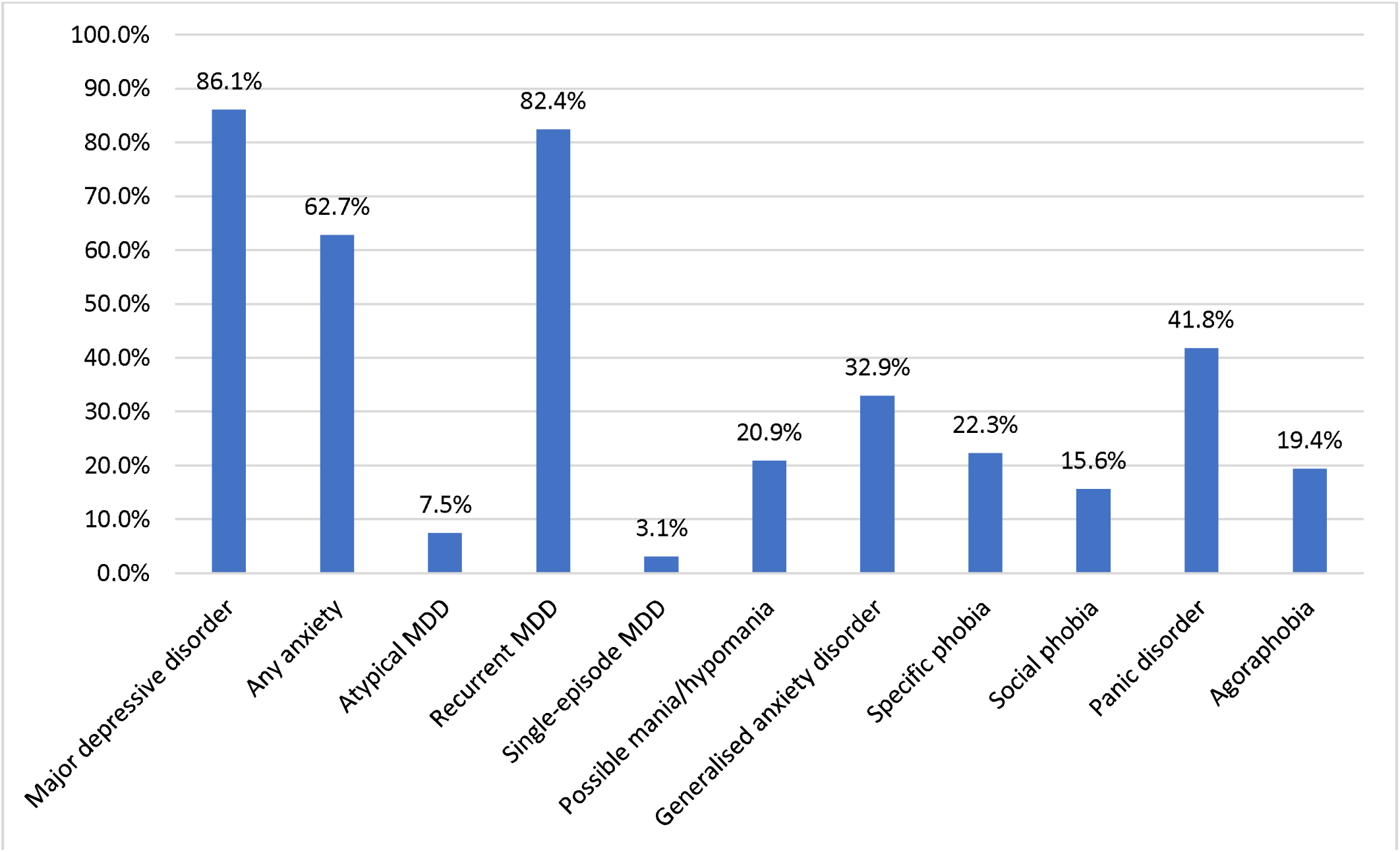
Prevalence of probable psychiatric disorders in the GLAD sample Percentages refer to the proportion of participants who meet cut-off criteria for the specified disorder as defined by the DSM-5 (American Psychiatric Association, 2013) out of the total. Possible mania was assessed by the Mood Disorder Questionnaire (MDQ) (Hirschfeld, 2002).

Retrospective reports of age of onset indicate a young average onset for the lifetime measures of major depressive disorder, specific phobia, social phobia, panic disorder, and agoraphobia (Table 4). Average age of onset across the anxiety disorders was ∼15. Age of onset was not measured for generalised anxiety disorder but will be added to the questionnaire in March 2019.

**Table 3.**
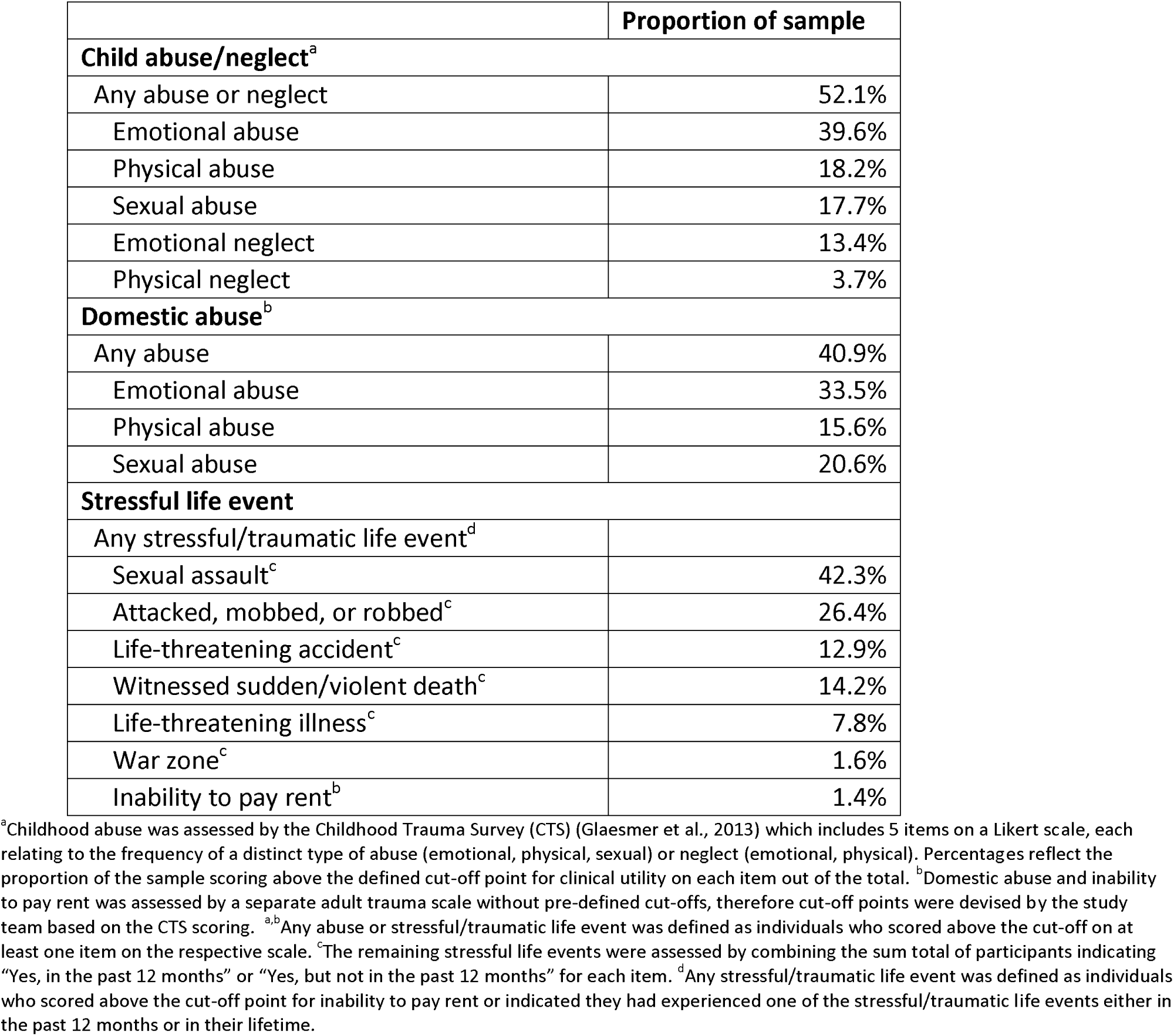
Proportion of the GLAD sample reporting previous child abuse/neglect, domestic abuse, or traumatic life events

|  | Proportion of sample |
| --- | --- |
| **Child abuse/neglect<sup>a</sup>** |  |
| Any abuse or neglect | 52.1% |
| Emotional abuse | 39.6% |
| Physical abuse | 18.2% |
| Sexual abuse | 17.7% |
| Emotional neglect | 13.4% |
| Physical neglect | 3.7% |
| **Domestic abuse<sup>b</sup>** |  |
| Any abuse | 40.9% |
| Emotional abuse | 33.5% |
| Physical abuse | 15.6% |
| Sexual abuse | 20.6% |
| **Stressful life event** |  |
| Any stressful/traumatic life event<sup>d</sup> |  |
| Sexual assault<sup>c</sup> | 42.3% |
| Attacked, mobbed, or robbed<sup>c</sup> | 26.4% |
| Life-threatening accident<sup>c</sup> | 12.9% |
| Witnessed sudden/violent death<sup>c</sup> | 14.2% |
| Life-threatening illness<sup>c</sup> | 7.8% |
| War zone<sup>c</sup> | 1.6% |
| Inability to pay rent<sup>b</sup> | 1.4% |

<sup>a</sup>Childhood abuse was assessed by the Childhood Trauma Survey (CTS) (Glaesmer et al., 2013) which includes 5 items on a Likert scale, each relating to the frequency of a distinct type of abuse (emotional, physical, sexual) or neglect (emotional, physical). Percentages reflect the proportion of the sample scoring above the defined cut-off point for clinical utility on each item out of the total. <sup>b</sup>Domestic abuse and inability to pay rent was assessed by a separate adult trauma scale without pre-defined cut-offs, therefore cut-off points were devised by the study team based on the CTS scoring. <sup>a,b</sup>Any abuse or stressful/traumatic life event was defined as individuals who scored above the cut-off on at least one item on the respective scale. <sup>c</sup>The remaining stressful life events were assessed by combining the sum total of participants indicating "Yes, in the past 12 months" or "Yes, but not in the past 12 months" for each item. <sup>d</sup>Any stressful/traumatic life event was defined as individuals who scored above the cut-off point for inability to pay rent or indicated they had experienced one of the stressful/traumatic life events either in the past 12 months or in their lifetime.

**Table 4.** Age of onset in GLAD sample for major depressive disorder, specific phobias, social phobia, panic disorder, and agoraphobia

|  | Median | Mean | SD |
| --- | --- | --- | --- |
| Major depressive disorder | 16 | 18.32 | 8.57 |
| Specific phobia | 10 | 11.07 | 8.13 |
| Social phobia | 12 | 13.52 | 8.09 |
| Panic disorder | 17 | 18.74 | 9.87 |
| Agoraphobia | 15 | 16.49 | 9.54 |

Measures of current symptoms of psychopathology included in the questionnaire demonstrate high rates of current clinical presentation of major depressive disorder and generalised anxiety disorder, with high levels of functional impairment (Table 5).

**Table 5.** Current symptoms of psychopathology and impairment in the GLAD sample.

|  | Mean | Range | SD | Proportion Clinical Severity |  |
| --- | --- | --- | --- | --- | --- |
| Major depressive disorder<sup>a</sup> | 11.08 | 0 - 27 | 6.866 | Clinical (total) | 80.5% |
|  |  |  |  | Mild (5 - 9) | 26.3% |
|  |  |  |  | Moderate (10 - 14) | 23.2% |
|  |  |  |  | Moderately severe (15 - 19) | 17.0% |
|  |  |  |  | Severe (20+) | 14.0% |
| Generalised anxiety disorder<sup>b</sup> | 9.02 | 0 - 21 | 5.987 | Clinical (total) | 73.5% |
|  |  |  |  | Mild (5 - 9) | 31.0% |
|  |  |  |  | Moderate (10 - 14) | 20.9% |
|  |  |  |  | Severe (15+) | 21.6% |
| Post-traumatic stress disorder<sup>c</sup> | 15.35 | 0-30 | 15.35 | Clinical (14+) | 56.5% |
| Personality disorder<sup>d</sup> | 7.98 | 0 - 25 | 3.586 | Clinical (total) | 51.8% |
|  |  |  |  | Mild (8 - 9) | 21.8% |
|  |  |  |  | Moderate (10+) | 30.0% |
| Alcohol use<sup>e</sup> | 7.15 | 0 - 40 | 6.392 | At-risk (total) | 37.7% |
|  |  |  |  | Increasing risk (8 - 15) | 26.6% |
|  |  |  |  | Higher risk (16 - 19) | 5.4% |
|  |  |  |  | Possible dependence (20+) | 5.8% |
| Functional Impairment<sup>f</sup> | 16.35 | 0-40 | 8.948 | Clinical (total) | 76.1% |
|  |  |  |  | Significant impairment (10 - 20) | 44.3% |
|  |  |  |  | Severe impairment (21+) | 31.8% |

Percentages refer to the proportion of participants out of the total sample who meet cut-off criteria for the specified disorder (as defined by the measures outlined below). <sup>a</sup> Criteria met for major depressive disorder on PHQ-9 (Kroenke et al., 2001) <sup>b</sup> Criteria met for generalised anxiety disorder on GAD-7 (Spitzer et al., 2006) <sup>c</sup> Criteria met for PTSD on PCL-6 (Lang et al., 2012) in the past month <sup>d</sup> Criteria met for a personality disorder on SASPD (Olajide et al., 2018) <sup>e</sup> Criteria met for at-risk alcohol dependence on AUDIT (Saunders et al., 1993) in the past year <sup>f</sup> Criteria met for functional impairment on WSAS (Mundt et al., 2002)

### Media Campaign

In order to assess the success of the media campaign, participants were asked to report how they found out about the GLAD Study at the end of the sign-up questionnaire. Of the 12,714 participants that completed the questionnaire, the 3 most common ways of hearing about the study were through Facebook, newspaper, and Twitter (SM 4).

The effectiveness of each recruitment strategy varied by age (see Figure 7). Of participants aged 16-29, the majority learned about the study through social media (Facebook, Twitter, Instagram, search engine), with only 12% receiving study information through traditional media (TV, radio, newspaper, online tabloid). Participants aged 30-49 also primarily learned about the study through social media, but 22% within that range learned about the study through traditional media. Social media was less effective in reaching individuals aged 50+, with traditional media being the primary means of recruitment above that age.

**Figure 7.**
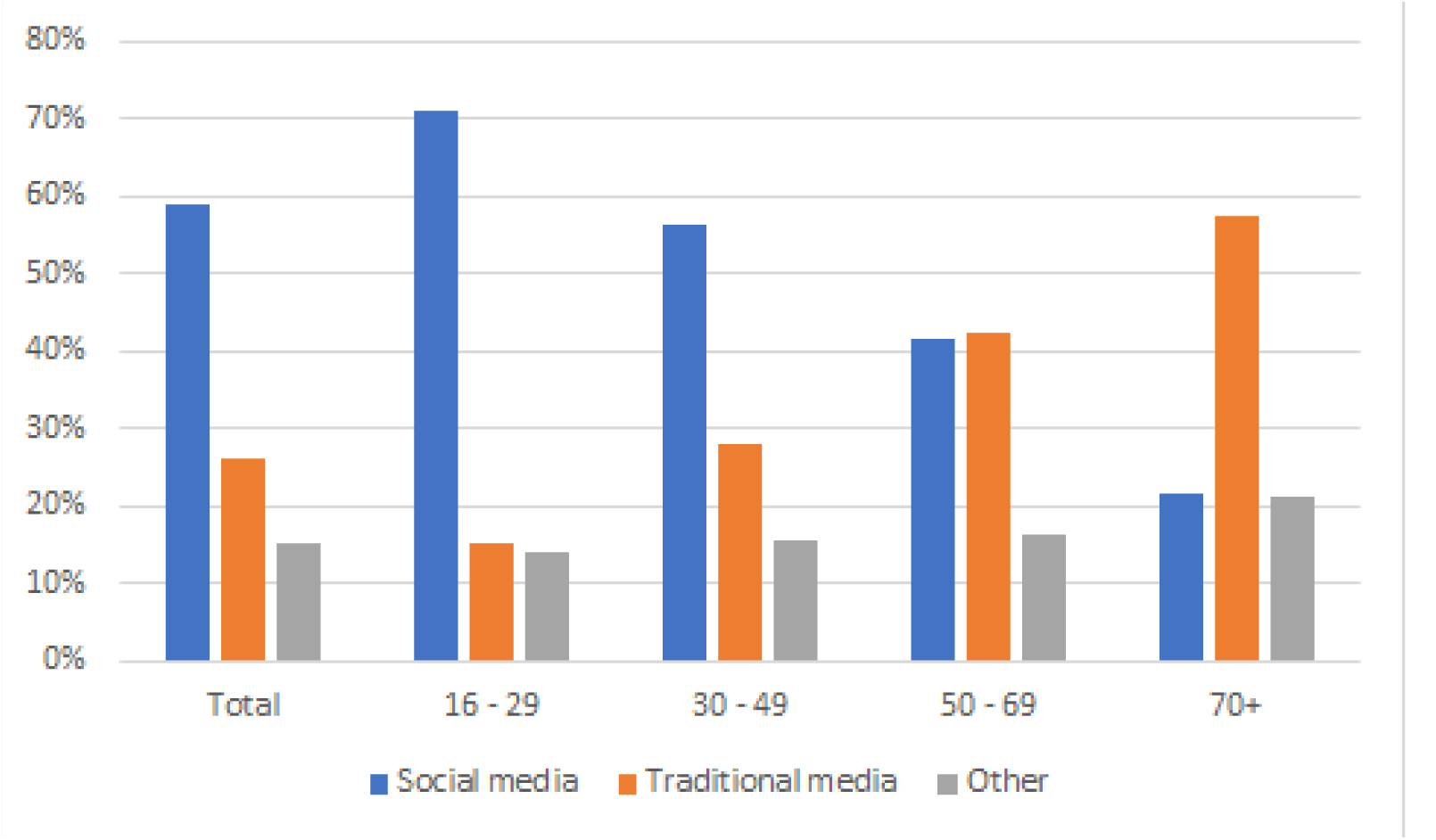
Media campaign drivers to complete questionnaires, by age group Participants were prompted to select all relevant responses to the question “How did you hear about the GLAD Study?” Social media refers to Facebook, Twitter, Instagram, bloggers, and search engines. Traditional media refers to radio, TV, newspaper, and online tabloids. All remaining responses are included in the Other category.

## Discussion

Overall, the GLAD Study represents a very large and comprehensive study of anxiety and depression and avaluable resource for future research. By achieving our goal of recruiting 40,000 participants with a lifetime occurrence of one of these disorders into the NIHR Mental Health BioResource, the study will collect detailed, homogenous phenotype and genotype data, thereby increasing power for genetic analyses. Importantly, the recontactable nature of the GLAD means that researchers will be able to conduct new studies integrating psychological and genetic data.

External researchers will be able to apply for access to anonymised data and will have the opportunity to recontact participants for additional data collection. This offers the prospect of a wide range of future recall studies, including clinical trials, observational studies, neuroimaging, and experimental medicine. Recruitment for recall studies could also be targeted to participants with phenotypes or genotypes of interest by utilising GLAD data to screen for specific eligibility criteria, thereby simplifying and expediting the recruitment process for future studies. Furthermore, measures were in part selected to match other cohorts and facilitate meta-analysis with samples such as the UK Biobank, Generation Scotland, and the Australian Genetics of Depression Study, although demographic differences between the samples would need to be taken into account (e.g. age differences between GLAD and UK Biobank participants). We propose that the GLAD data could be useful for stratifying cases into distinct phenotypes of depression and anxiety to reduce heterogeneity, a strategy which has been shown to increase the power of genetic analyses to detect significant effects (CONVERGE consortium, 2015; Power et al., 2017; Verduijn et al., 2017).

Initial descriptive analyses reveal that the current GLAD sample does not reflect the demographics of general population of England. The limited ethnic diversity within the sample is not representative, with 94% of participants being white compared to 85% of the population. Similarly, although common mental disorders are more than twice as common in young women compared to young men, and 1.5 times more common in women overall (NHS Digital, 2016), the GLAD sample remains disproportionately female. Finally, the study sample report roughly double the proportion of participants with a university degree compared to the English population.

The GLAD sample as a whole also shows severe psychopathology. Our lifetime questionnaire diagnostic algorithm indicated that the majority of the sample have recurrent depression, and 20.9% report possible mania/hypomania. Over half the sample screen positively for a lifetime anxiety disorder, with panic disorder being the most common. Participants also show substantial comorbidity, in particular for GAD cases reporting a concurrent diagnosis of MDD. Our results show significantly higher comorbidity rates for GAD cases compared to a previous epidemiological study whereby 62.4% of GAD cases had a lifetime occurrence of MDD (Judd et al., 1998). However, this is likely further indication of the sample’s severity, reflecting previous reports of higher rates of comorbid anxiety in patients with recurrent lifetime depression (Boland, Keller, Gotlib, & Hammen, 2002; Moffitt et al., 2007). In terms of current symptoms, GLAD participants report moderate symptoms of depression, mild to moderate symptoms of anxiety, and significant functional impairment, indicating poor work and social adjustment. Finally, it was interesting to note that 51.8% of participants screened positive for mild personality disorder.

Severity levels are also demonstrated by the 95% of GLAD participants who report receiving treatment for depression or anxiety, although the study is open to individuals regardless of treatment receipt. In Western countries, it is estimated that between one-thirds and one half of patients with depression or anxiety disorders do not receive a diagnosis and/or treatment (D. Kessler, Bennewith, Lewis, & Sharp, 2002), a group not represented in our sample. However, even in those patients receiving treatment, recent research suggests that only 20% get minimally adequate treatment (Thornicroft et al., 2017), meaning that follow-up research on treatment efficacy in the GLAD sample would be highly valuable.

An unanticipated benefit of the campaign was the interest it generated from UK NHS sites around the country. Over 45 NHS sites, including Trusts and GP practices, contacted the study team with interest in supporting recruitment, and many had learned about the study from the media campaign. The study was also adopted onto the NIHR Clinical Research Network (CRN) Portfolio which provides support and funding for NHS organisations that are involved in research. The combined approach of both general and clinical recruitment reaches a wider demographic of participants than either strategy alone. Patients recruited through clinics can also be assisted in signing up by a local clinician or research team to provide support throughout the process if needed.

In addition to collecting a large amount of data, the GLAD study represents a template for future online studies. The media campaign proved highly effective in recruiting a large number of participants in a short amount of time and demonstrates the success of such a broad outreach approach. Caution must be taken when interpreting the success of the different media campaign strategies, given the prolonged use of social media promotions in comparison to traditional media outlets; nonetheless social media was the most effective strategy for recruiting individuals under 50. Recruiting younger participants not only reflects the young average age of onset of anxiety and depression, approximately 11 (Bandelow & Michaelis, 2015) and 24 (Christie et al., 1988; R. C. Kessler et al., 2003) respectively, but also helps to facilitate long-term follow up and recontact.

### Limitations

As previously mentioned, the current demographics of study participants who completed the questionnaire are not representative of the population of England, indicating a likely self-referral bias. Results from analyses of the current GLAD sample therefore may not be generalisable to the whole population. Smaller studies could overcome these biases by selectively over-sampling males and individuals at the lower end of educational attainment to be more representative of the population. However, efforts are being taken to recruit a wider demographic of participants into the study.

Specifically, we will develop a targeted social media campaign to reach young men and collaborate with young male influencers to appeal to that audience. We will additionally prepare a future media campaign that focuses on depression and anxiety separately, with wording specific to each disorder, to attempt to reduce the high comorbidity rates in the sample. Furthermore, we are collaborating with local branches of the charity Mind. These are typically placed within the community and provide a range of mental health support services. Branches will be displaying posters and leaflets on site, posting on social media, and answering questions or providing assistance to potential participants interested in signing up. Recruitment through NHS services and the availability of local research teams within those sites will also help reach the general population and recruit individuals with a wider range of educational attainment.

Of particular importance, we are working with Black, Asian, and minority ethnic charities and influencers to conduct additional user testing to understand the barriers to participating for non-white individuals and increase outreach to diverse communities. Previous genetic studies have involved primarily individuals of white European ancestry (Haga, 2010), and findings from these studies may not apply to individuals of non-white descent. By actively recruiting a diverse sample, our objective is to additionally facilitate research on typically underrepresented groups.

Another challenge of the study design is the drop-out rates following recruitment. At each stage of the sign-up process, a substantial number of participants did not complete the next step despite multiple reminders. Unfortunately, we are unable to assess the demographics of participants who do not complete the sign-up questionnaire, but it is possible that the drop-outs are non-random and represent a certain phenotype, such as individuals with more severe symptoms or lower health literacy.

## Conclusion

The GLAD study offers a recontactable resource of participants with a lifetime occurrence of depression and anxiety disorders to facilitate future health research. The online study design and media recruitment strategy were effective in recruiting a large number of individuals with these disorders into the NIHR Mental Health BioResource. Recruitment is ongoing with the goal of completing recruitment of 40,000 individuals, making this the largest single study of depression and anxiety. We hope that this paper will not only demonstrate the effectiveness of the study methodology but will also raise awareness of the availability of this cohort to researchers in the field and promote future collaboration.

## Data Availability

This is a cohort description paper.

## Abbreviations

SNP: single nucleotide polymorphism
GLAD: Genetic Links to Anxiety and Depression
NIHR: National Institute for Health Research
IAPT: Improving Access to Psychological Therapies
SLaM: South London and Maudsley
NHS: National Health Service
SURE: Service User Research Enterprise
FAST-R: Feasibility and Acceptability Support Team for Researchers
PR: public relations
SOPs: standard operating procedures
CRN: Clinical Research Networks
GP: general practitioner
CCG: Clinical Commissioning Group

## Acknowledgements

We gratefully acknowledge the participation of all GLAD Study volunteers, and thank the NIHR Mental Health BioResource staff for their help with volunteer recruitment. This study presents independent research funded by the National Institute for Health Research (NIHR) Biomedical Research Centre at South London and Maudsley NHS Foundation Trust and King’s College London. Further information can be found at http://brc.slam.nhs.uk/about/core-facilities/bioresource.

## Notes

### Competing Interest Statement

The authors have declared no competing interest.

### Funding Statement

UK NIHR

### Author Declarations

All relevant ethical guidelines have been followed and any necessary IRB and/or ethics committee approvals have been obtained.

Any clinical trials involved have been registered with an ICMJE-approved registry such as ClinicalTrials.gov and the trial ID is included in the manuscript.

